# Detecting Contradictory Evidence on Drug-Disease Associations at Scale

**DOI:** 10.64898/2026.11.11.26367856

**Authors:** Mengfei Lan, Joe Menke, Shiwei Liu, Zhiyong Lu, Halil Kilicoglu

## Abstract

Contradictory claims about drug efficacy across randomized controlled trials (RCTs) complicate evidence-based medicine, yet existing automated methods classify isolated sentence pairs and do not explain why studies disagree. We present **DrugCon**, a large language model (LLM)-based framework that extracts study conclusions from RCT abstracts, links these claims when they concern the same drug–disease association, detects contradictions with counterfactual reasoning, and explains them using a taxonomy of ten study-level factors such as population, dosage, and outcome measures. Applying **DrugCon** to 224,756 RCT abstracts, we built DrugCon-Bench, an exhaustively annotated benchmark of 542 contradictory claim pairs from 819 abstracts. Most contradictions involve more than one study-level factor (75.5%). The most common are differences in outcome measures (64.0%), populations (51.3%), and dosage or exposure duration (35.6%). **DrugCon** achieves an F_1_ score of 0.744 for contradiction detection, outperforming rule-based, supervised, and natural language inference baselines, and an F_1_ of 0.482 for identifying explanatory study factors. **DrugCon** enables scalable and explainable analysis of contradictory clinical evidence.

## 1 Introduction

Biomedical literature is a major source of scientific evidence for understanding disease mechanisms, evaluating treatment effectiveness, and guiding clinical decision-making. Among biomedical evidence sources, randomized controlled trials (RCTs) are considered the gold standard for evaluating drug efficacy and establishing reliable drug–disease associations due to their ability to minimize bias and support causal inference (Pallari and Lewison 2019; Larson and White 2015; Naci and Ioannidis 2015; Pitrou et al. 2009). With approximately 700,000 RCT papers indexed in PubMed between 1990 and 2020 (Zhao et al. 2022), RCT literature provides a rich resource for evidence synthesis (Manion et al. 2024), clinical decision support (Devereaux and Yusuf 2003; Chen et al. 2008), and drug repurposing (Liu et al. 2021). In particular, large-scale analysis of RCT literature has become an important approach for identifying potential drug–disease associations and supporting drug repurposing efforts (Makhervaks et al. 2023).

However, findings reported across RCT studies could be contradictory. Analyses of highly cited clinical studies have shown that some influential findings are later contradicted by subsequent investigations (Ioannidis 2005a; Prasad et al. 2013). Such contradictions may arise from differences in study populations, experimental settings, drug dosages, outcome measures, or methodological design (Ioannidis 2005a; Rosemblat et al. 2019), suggesting that understanding contradictions requires not only identifying contradictory evidence but also interpreting the study differences from the contexts. Figure 1 illustrates an example in which two studies examining the same intervention (indomethacin for pancreatitis) reach different conclusions. This contradiction may be explained by differences in study population, as the studies were conducted in the USA vs. China, which can differ in pancreatitis risk factors, drug metabolism genetics, and clinical practice patterns (Rothwell 2005).

**Fig. 1.**
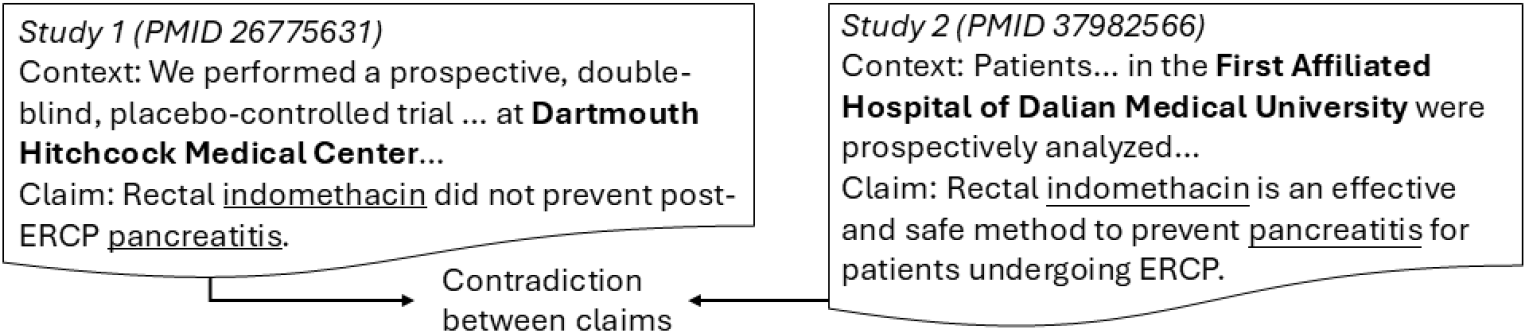
Illustration of contradiction in scientific papers, where studies exploring the same drug intervention (indomethacin for pancreatitis) while making different suggestions.

Identifying and explaining contradictory drug-disease associations across RCTs is important for evidence-based decision-making. For drug repurposing research, systematically identifying and interpreting contradictory findings across RCTs can improve the reliability of the evidence base used to generate drug repurposing hypotheses (Makhervaks et al. 2023). For clinicians and policymakers, understanding and reconciling contradictory drug-disease associations is essential for producing reliable evidence syntheses and meta-analyses (Tsafnat et al. 2014; Marshall and Wallace 2016). Vitamin D supplementation for childhood asthma illustrates the challenge of contradictory evidence: a study reports that vitamin D supplementation improves asthma control in children (PMID 26841365), while another finds that it does not improve asthma control in children (PMID 33522698). However, the two studies differ in dosage and duration (800 IU/day for 2 months vs. 2000 IU/day for 12 weeks) and outcome measures (physician-assessed GINA asthma control levels vs. childhood asthma control test scores), which may explain their different findings. Without such explanations, contradictory evidence is difficult to synthesize, which motivates automated approaches to identify and explain contradictions at scale (Ioannidis 2016; Sosa et al. 2023).

Despite the importance of understanding contradictions, there is currently no scalable framework for systematically identifying and explaining contradictions across the biomedical literature. Prior work on contradiction detection falls into three groups. First, rule-based methods detect contradictions from surface patterns, such as polarity differences between claims (Kim et al. 2006; Sarafraz 2012) or SemRep triples with the same subject and object but opposing predicates (e.g., *treats* vs. *causes*) (Kilicoglu et al. 2020; Rosemblat et al. 2019). To the extent that these methods capture contradictions at all, they are limited to explicit oppositions and fail to detect those expressed implicitly. Second, supervised methods train classifiers on curated datasets of contradictory claim pairs (Alamri and Stevenson 2016; Tawfik and Spruit 2018; Yazi et al. 2021; Makhervaks et al. 2023), but depend on their training data and cannot generalize to new clinical topics. Third, natural language inference (NLI) methods treat contradiction detection as an entailment classification task using models trained on NLI corpora such as MNLI and MedNLI (Williams et al. 2018; Romanov and Shivade 2018), but are trained on short sentence pairs and miss contradictions that depend on the claim context (Section 2.3). All three groups of methods operate on isolated sentence pairs and approach contradiction detection as a binary classification task, but do not explore how to contextualize the source of contradictions. Many apparent contradictions in biomedical findings are not true inconsistencies, but instead arise from differences in study design, population, or outcome definitions (Rosemblat et al. 2019). Therefore, detecting contradictions is insufficient for evidence synthesis, where understanding the difference of two study context is important to interpret the evidence.

To address these challenges, we propose **DrugCon**, an LLM-based framework for detecting and explaining contradictory drug–disease association claims in RCT documents at scale (224,756 abstracts). Following the definition of Rosemblat et al. (2019), we focus on apparent contradictions: claims that appear inconsistent or opposing, although the opposition can often be explained by differences in study conditions, such as population, dosage, outcome measures, or study design. This contrasts with genuine contradictions, logical inconsistencies in which one claim must be false, which are rare in the life sciences (Rosemblat et al. 2019; Ioannidis 2005a). **DrugCon** focuses on apparent contradictions because interpreting them requires reasoning over study context rather than comparing claim statements alone. As illustrated in Figure 2, **DrugCon** extracts claims from RCT abstracts, identifies semantically related claim pairs, and determines whether the claims present contradictory findings. Unlike existing methods, **DrugCon** generates explanations for contradictions by leveraging full abstract context. To support contradiction explanation, we adapt and extend a taxonomy of ten study-level factors (e.g., population, dosage, and outcome measures) (Rosemblat et al. 2019) that systematically capture sources of variation across studies. These explanations also allow experts to verify and prioritize the detected contradictions without re-reading each pair of abstracts. By organizing contextual evidence according to this taxonomy, **DrugCon** identifies study differences that may explain contradictory findings.

**Fig. 2.**
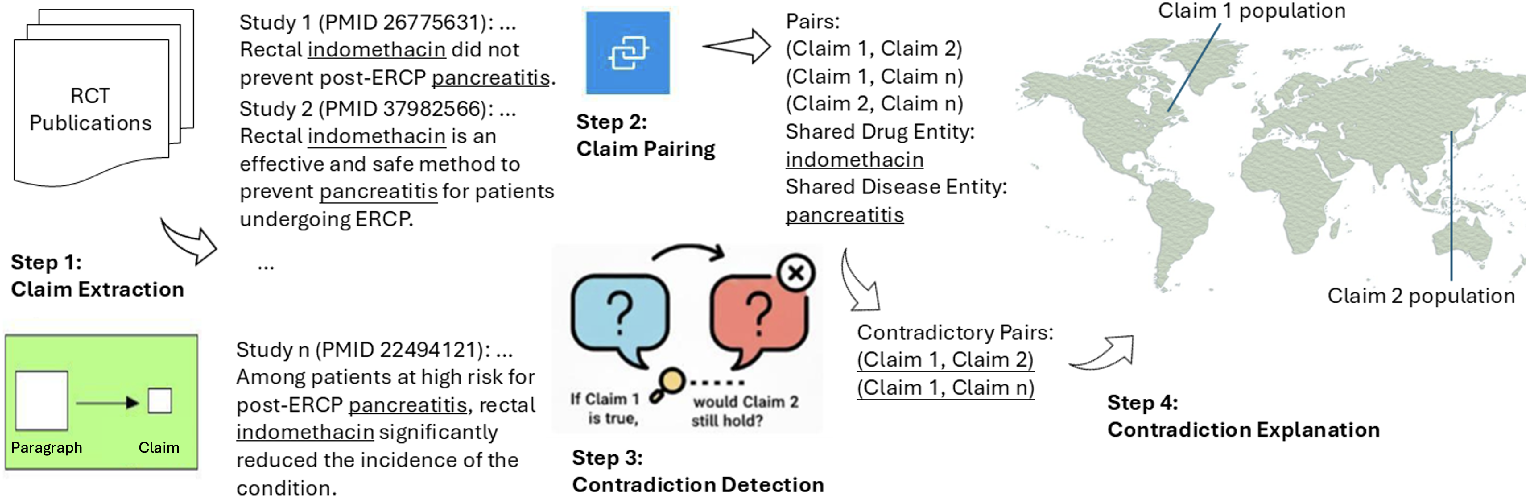
_Overview of_ **_DrugCon_** _framework for drug-disease contradiction detection in large-scale RCT literature. The example illustrates a contradiction between PMID 26775631 and PMID 37982566, explained by the *population* study factor._

To evaluate the proposed framework, we apply **DrugCon** to 224,756 RCT abstracts and conduct expert annotation to assess the quality of contradiction detection and interpretation. Based on these annotations, we construct DrugCon-Bench, a benchmark dataset containing exhaustively annotated contradictory drug–disease claim pairs from a subset of RCT abstracts, enabling reliable evaluation of contradiction detection and study-factor based explanations. To further evaluate the generalizability of **DrugCon**, we conduct experiments on an existing benchmark, ManConCorpus (Alamri and Stevenson 2016), which contains contradictory claim annotations from publications on cardiovascular diseases.

Our contributions are as follows:

- We introduce **DrugCon**, an LLM-based framework that not only detects but also explains contradictory drug–disease associations in RCTs, addressing prior limitations of sentence-level contradiction detection, lack of explanations, and poor generalization beyond supervised datasets.
- We adapt and extend a taxonomy of ten study-level factors (e.g., study design,

population, and outcome measures) that captures differences between study contexts that may lead to contradictions and enables **DrugCon** to generate structured explanations using the full abstract context.

- We construct DrugCon-Bench, a benchmark dataset of 542 expert-annotated contradictory claim pairs, where annotators labeled each pair with factors in the taxonomy to explain study differences underlying the contradiction, covering all pair-wise contradictions from 819 RCT abstracts.
- Through extensive experiments on DrugCon-Bench and the external ManConCorpus dataset, we demonstrate that **DrugCon** effectively identifies contradictory evidence and leverages contextual information to explain the underlying study differences that may lead to disagreement.

## 2 Methods

In this section, we present **DrugCon**, our contradictory evidence detection framework. We frame literature-based contradiction detection for drug–disease associations as a hybrid task combining semantic claim linking and conditional text generation. Given a large corpus of biomedical texts, the **DrugCon** framework consists of four stages: (1) *claim extraction* -extract the set of claims made in each document; (2) *claim pairing* - identify the claim pairs that mention the same drug-disease association and are semantically similar based on dense vector representations of the claims; (3) *contradiction detection* - use conditional generation and reasoning abilities of LLMs to detect contradictions within the linked claim pairs; (4) *contradiction explanation* - explain the contradictions in the claims by reasoning about the study-level factor difference from the contexts of the claims. To support the evaluation of the framework outputs, we conduct (5) *ground truth annotation*: establishing human-annotated ground truth to evaluate the quality of contradiction detection and explanation outputs. We finally conduct (6) *ablation study* on the choice of backbone LLM and prompt design. Figure 2 presents an overview of **DrugCon** framework. The detailed procedure for each stage is described below.

### 2.1 Claim Extraction

RCT abstracts are typically organized into structured sections (e.g., background, methods, results, and conclusions) (Schulz et al. 2010). The CONSORT extension for abstracts (Hopewell et al. 2008) recommends that the Conclusions provide a general interpretation of the trial’s results, making conclusion sentences the most appropriate source for authors’ claims. In line with this conclusion definition, we focus on conclusion sentences as the primary scientific claims of RCT abstracts. The models dedicated to claim extraction (Achakulvisut et al. 2019; Wright et al. 2022) identify verifiable or checkworthy statements as claims, but do not distinguish between result-level observations and the author’s conclusions. For example, in an RCT abstract, both “the drug reduced blood pressure by 10 mmHg (p *<* 0.05)” and “these findings suggest that the drug is effective for treating hypertension” are verifiable and checkworthy statements and classified as claims by the models dedicated to claim extraction, but only the latter represents the author’s overall conclusion about the drug-disease association. Therefore, we leveraged a model proposed by an existing work (Lan et al. 2024), which automatically classifies abstract sentences into rhetorical categories (background, objective, methods, results, and conclusions) that achieves state-of-the-art performance on conclusion sentence identification (F_1_=0.92).

### 2.2 Claim Pairing

Based on the extracted claims, we construct pairs that discuss the same drug–disease association. This process follows an information retrieval paradigm, where the goal is to identify all claims corresponding to a single drug-disease association. In biomedical retrieval, sparse methods (e.g., BM25 (Robertson et al. 2009)) are efficient but limited to surface-level matching (Guo et al. 2022), while dense methods (e.g., MedCPT (Jin et al. 2023), BMRetriever (Xu et al. 2024), and BioHiCL (Lan et al. 2026)) better capture semantic similarity at higher computational cost. To balance these trade-offs, we adopt a two-stage coarse-to-fine framework: sparse retrieval first generates candidate claim pairs, which are then refined using dense retrieval for improved semantic alignment. The details are as follows:

### Entity Pairing

We first identify topic-related claims by linking those that share the same drug–disease association. To achieve this, we use PubTator 3 (Wei et al. 2024), a widely used biomedical text-mining tool that achieves high performance on drug and disease recognition, to recognize and normalize drug and disease entities in each claim to MeSH identifiers. Claims (the extracted conclusion sentences) are then paired if they contain at least one overlapping normalized drug and one overlapping normalized disease, indicating they refer to the same drug–disease pair.

### Dense Pairing

Entity pairing based on shared drug–disease entities may still link claims that discuss different aspects of the association and are not semantically comparable. To filter such cases, we apply a dense retrieval model to assess semantic similarity between claims. Specifically, we use BioHiCL-Base (Lan et al. 2026), a biomedical dense retriever trained with hierarchical MeSH supervision to capture fine-grained semantics. We encode each claim into a vector representation and compute cosine similarity for each candidate pair. Pairs with similarity above 0.7 are retained. The threshold 0.7 is determined empirically by manually checking claim pairs at different similarity thresholds and selecting the value that best balances retaining comparable pairs while removing unrelated matches.

### 2.3 Contradiction Detection

Given a pair of claims referring to the same drug–disease association, we then determine whether they contain contradictory information. We leverage LLMs for their conditional generation capabilities and contextual reasoning, designing prompts that instruct the LLMs to perform contradiction detection.

A key challenge in contradiction detection is that contradictions are often partial, arising at the level of specific subclaims rather than entire claim sentences. For example, consider the following pair of claims:

- Claim 1: *“****Cariprazine*** *1.5 mg/day significantly reduced* ***depressive symptoms*** *in adults with bipolar I depression compared to placebo, but differences were not significant for* ***cariprazine*** *3.0 mg/day.”* (PMC7318333)
- Claim 2: *“****Cariprazine*** *1.5–3 mg/d consistently improved* ***depressive symptoms*** *in all patient subgroups without regard to differences in baseline demographic and clinical characteristics.”* (PMC7846289)

Here, Claim 1 reports that the 1.5 mg/day dose reduces depressive symptoms, whereas the 3.0 mg/day dose does not. Claim 2, in contrast, states that both 1.5–3 mg/day doses consistently improve depressive symptoms across patient subgroups. Thus, the two claims agree on the effectiveness of the 1.5 mg/day dose but disagree on the effectiveness of the 3.0 mg/day dose. This example illustrates that a single scientific claim can contain multiple embedded subclaims, some of which may align with another claim while others contradict it. This requires models to identify which subclaim of two complex claim sentences should be compared. Prior work in natural language inference and fact-checking (Zhu et al. 2023; Zhao et al. 2024; Li et al. 2024) addresses this issue using the decompose-then-verify framework, which breaks claims into smaller units of subclaims for fine-grained comparison (Min et al. 2023; Song et al. 2024; Chern et al. 2023). However, recent studies show that decomposition does not improve reasoning over complex scientific text, as it can introduce additional variability that propagates to downstream tasks (Hu et al. 2025).

We avoid explicit claim decomposition and instead adopt a counterfactual reasoning approach that preserves the full claim context (Goyal et al. 2019). Specifically, we prompt LLMs to evaluate whether one claim would still hold if the other were assumed to be true:

- If Claim 1 is true, would Claim 2 still hold?
- If Claim 2 is true, would Claim 1 still hold?

For the cariprazine example, assuming Claim 1 is true (3.0 mg/day is not effective), Claim 2’s assertion that 1.5–3 mg/day consistently improves symptoms no longer holds. Assuming Claim 2 is true, Claim 1’s null finding for 3.0 mg/day does not hold either. Both directions reveal an incompatibility, so the pair is labeled contradictory. A contradiction is identified if either direction reveals the incompatibility. If assuming that one claim invalidates the other, the two claims cannot simultaneously hold, which is consistent with the definition of contradiction in natural language inference (MacCartney and Manning 2009). This counterfactual prompting strategy enables implicit subclaim-level comparison while maintaining contextual coherence. The detailed prompt is provided in Appendix B.1. The LLM output is then processed to extract the binary contradiction label, and pairs labeled as contradictory are passed to the explanation step to identify the corresponding study-level factors (Section 2.4).

### 2.4 Contradiction Explanation

After identifying contradictory claim pairs, we use LLMs to explain the underlying reasons for the disagreement based on study-level factors in the full abstract context of each claim.

Existing research has shown that contradictions in biomedical texts often arise from differences in the contextual conditions under which claims are generated, rather than from genuine scientific contradictions. Ioannidis (2005b) argues that discrepancies across experimental results could stem from variations in study design, sample size, and statistical power, as well as from the natural evolution of scientific evidence. Van der Worp et al. (2010) present that interventions effective in animal models may fail in human studies due to species-specific biological and disease mechanism differences. Voils et al. (2012) further show that differences in dosage and exposure conditions can lead to different outcomes. Building on these insights, Rosemblat et al. (2019) propose a structured taxonomy of contextual factors to systematically characterize variation of study-level factors. However, previous contradiction detection approaches operate on isolated sentence or claim pairs. They cannot explain contradictions using study-level factors because the relevant contextual information, such as patient population, intervention details, outcome measures, and study design, is often distributed throughout the abstract rather than contained in the claim sentence alone.

We adopt and extend this taxonomy (Rosemblat et al. 2019) to characterize the context of drug-disease knowledge in RCTs. Specifically, we:

- Retain six categories: *Species*, *Population* (merged with *Associated condition*, since in RCTs, comorbidities are part of the participant characteristics that define who is enrolled, considering age, sex, and disease severity), *Dosage and exposure duration*, *Route or mode of administration*, *Evolving scientific evidence* (originally *New knowledge*), and *Known controversy*.
- Introduce four categories that capture trial-level context reported in RCT abstracts: *Outcome measures*, *Combined drug effect*, *Study design*, and *Ambiguous referent*.
- Exclude two categories that do not apply to drug–disease RCTs: *Idiosyncratic reaction*, which reflects individual responses rather than group-level trial findings; and *Endogenous/exogenous*, because trial interventions are always administered exogenously.

The developed taxonomy containing 10 study-level factors (e.g., population, dosage, route of administration, …) is presented in Table 1.

**Table 1:**
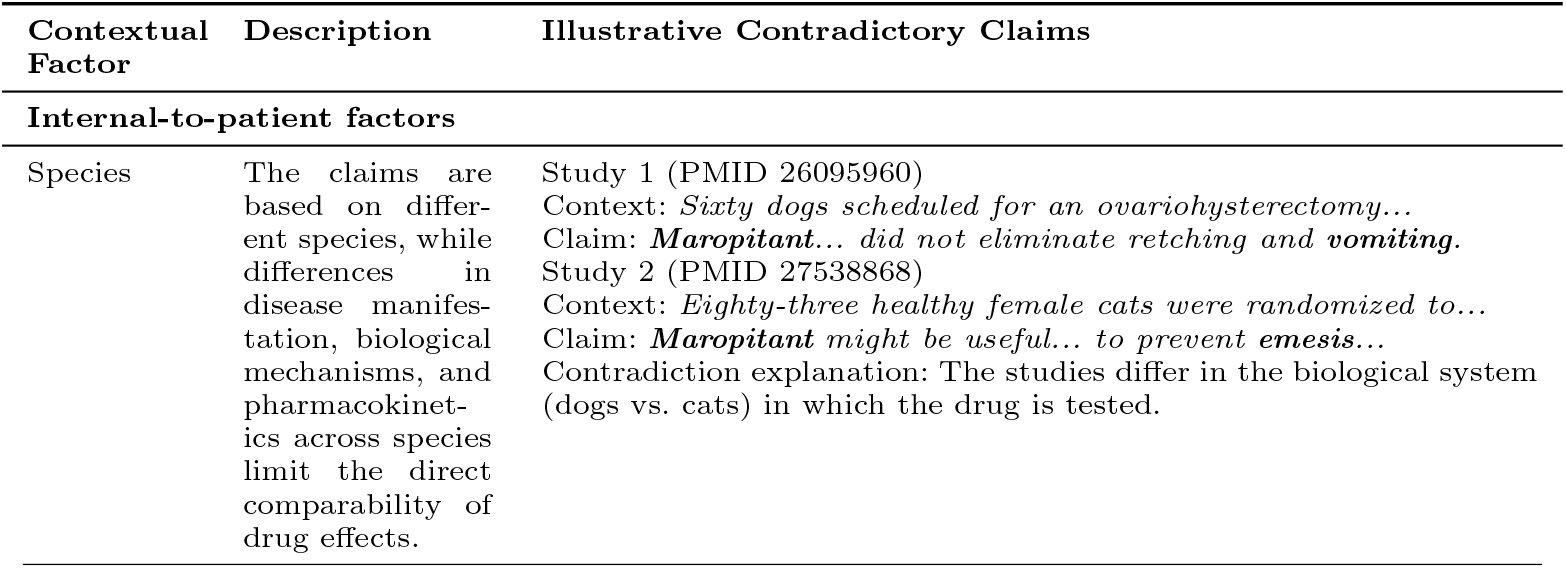
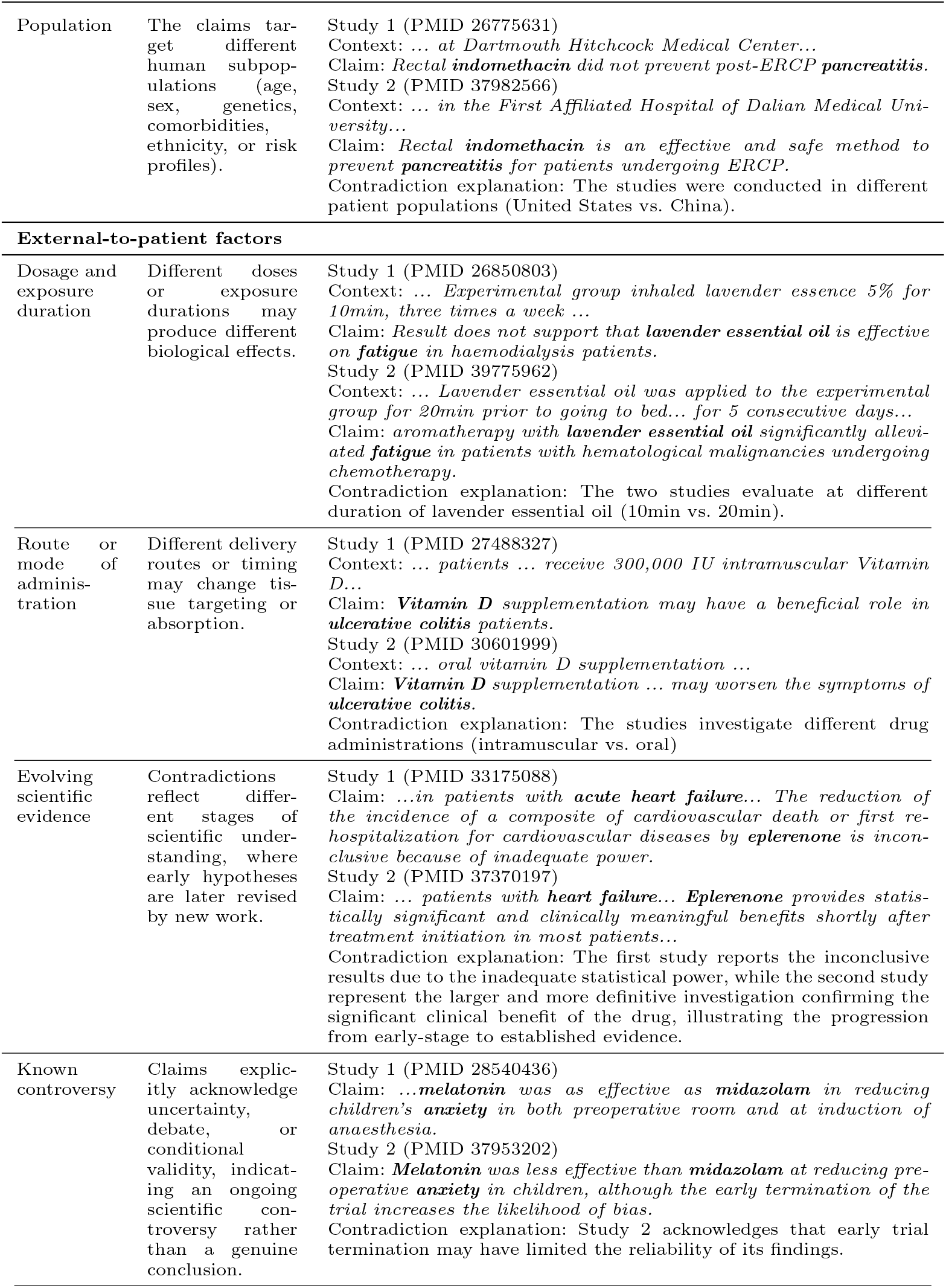
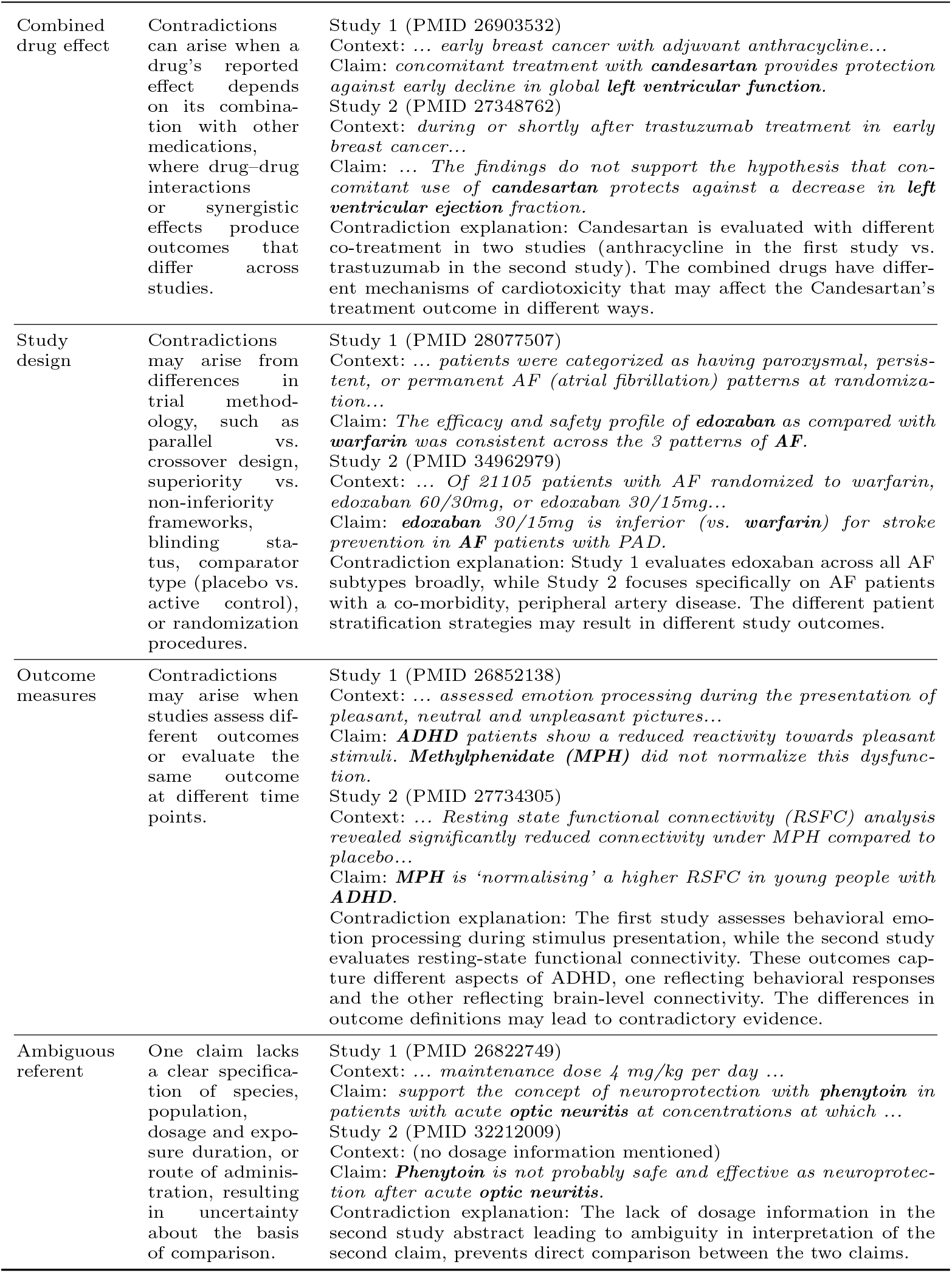
Taxonomy of contextual factors for explaining contradictory RCT findings, adapted and extended from Rosemblat et al. (2019). The column “Illustrative Contradictory Claims” presents example claim pairs whose contradictions can be explained by the corresponding study-level contextual factor. **Bold** highlights the key drug and disease entities in the claims.

| **Contextual Factor** | **Description** | **Illustrative Contradictory Claims** |
|---|---|---|
| **Internal-to-patient factors** |  |  |
| Species | The claims are based on differ-ent species, while differences in disease manifes-tation, biological mechanisms, and pharmacokinet-ics across species limit the direct comparability of drug effects. | Study 1 (PMID 26095960)<br>Context: *Sixty dogs scheduled for an ovariohysterectomy...*<br>Claim: ***Maropitant****... did not eliminate retching and* ***vomiting.***<br>Study 2 (PMID 27538868)<br>Context: *Eighty-three healthy female cats were randomized to...*<br>Claim: ***Maropitant*** *might be useful... to prevent* ***emesis****...*<br>Contradiction explanation: The studies differ in the biological system (dogs vs. cats) in which the drug is tested. |
| Population | The claims tar-get different human subpop-ulations (age, sex, genetics, comorbidities, ethnicity, or risk profiles). | Study 1 (PMID 26775631)<br>Context: *... at Dartmouth Hitchcock Medical Center...*<br>Claim: *Rectal* ***indomethacin*** *did not prevent post-ERCP* ***pancreatitis.***<br>Study 2 (PMID 37982566)<br>Context: *... in the First Affiliated Hospital of Dalian Medical Uni-versity...*<br>Claim: *Rectal* ***indomethacin*** *is an effective and safe method to prevent* ***pancreatitis*** *for patients undergoing ERCP.*<br>Contradiction explanation: The studies were conducted in different patient populations (United States vs. China). |
| **External-to-patient factors** |  |  |
| Dosage and exposure duration | Different doses or exposure durations may produce different biological effects. | Study 1 (PMID 26850803)<br>Context: *... Experimental group inhaled lavender essence 5% for 10min, three times a week ...*<br>Claim: *Result does not support that* ***lavender essential oil*** *is effective on* ***fatigue*** *in haemodialysis patients.*<br>Study 2 (PMID 39775962)<br>Context: *... Lavender essential oil was applied to the experimental group for 20min prior to going to bed... for 5 consecutive days...*<br>Claim: *aromatherapy with* ***lavender essential oil*** *significantly allevi-ated* ***fatigue*** *in patients with hematological malignancies undergoing chemotherapy.*<br>Contradiction explanation: The two studies evaluate at different duration of lavender essential oil (10min vs. 20min). |
| Route or mode of adminis-tration | Different delivery routes or timing may change tis-sue targeting or absorption. | Study 1 (PMID 27488327)<br>Context: *... patients ... receive 300,000 IU intramuscular Vitamin D...*<br>Claim: ***Vitamin D*** *supplementation may have a beneficial role in* ***ulcerative colitis*** *patients.*<br>Study 2 (PMID 30601999)<br>Context: *... oral vitamin D supplementation ...*<br>Claim: ***Vitamin D*** *supplementation ... may worsen the symptoms of* ***ulcerative colitis.***<br>Contradiction explanation: The studies investigate different drug administrations (intramuscular vs. oral) |
| Evolving scientific evidence | Contradictions reflect differ-ent stages of scientific under-standing, where early hypotheses are later revised by new work. | Study 1 (PMID 33175088)<br>Claim: *...in patients with* ***acute heart failure****... The reduction of the incidence of a composite of cardiovascular death or first re-hospitalization for cardiovascular diseases by* ***eplerenone*** *is incon-clusive because of inadequate power.*<br>Study 2 (PMID 37370197)<br>Claim: *... patients with* ***heart failure****...* ***Eplerenone*** *provides statis-tically significant and clinically meaningful benefits shortly after treatment initiation in most patients...*<br>Contradiction explanation: The first study reports the inconclusive results due to the inadequate statistical power, while the second study represent the larger and more definitive investigation confirming the significant clinical benefit of the drug, illustrating the progression from early-stage to established evidence. |
| Known controversy | Claims explic-itly acknowledge uncertainty, debate, or conditional validity, indicat-ing an ongoing scientific con-troversy rather than a genuine conclusion. | Study 1 (PMID 28540436)<br>Claim: *...****melatonin*** *was as effective as* ***midazolam*** *in reducing children’s* ***anxiety*** *in both preoperative room and at induction of anaesthesia.*<br>Study 2 (PMID 37953202)<br>Claim: ***Melatonin*** *was less effective than* ***midazolam*** *at reducing pre-operative* ***anxiety*** *in children, although the early termination of the trial increases the likelihood of bias.*<br>Contradiction explanation: Study 2 acknowledges that early trial termination may have limited the reliability of its findings. |
| Combined drug effect | Contradictions can arise when a drug’s reported effect depends on its combina-tion with other medications, where drug–drug interactions or synergistic effects produce outcomes that differ across studies. | Study 1 (PMID 26903532)<br>Context: *... early breast cancer with adjuvant anthracycline...*<br>Claim: *concomitant treatment with* ***candesartan*** *provides protection against early decline in global* ***left ventricular function.***<br>Study 2 (PMID 27348762)<br>Context: *during or shortly after trastuzumab treatment in early breast cancer...*<br>Claim: *... The findings do not support the hypothesis that con-comitant use of* ***candesartan*** *protects against a decrease in* ***left ventricular ejection*** *fraction.*<br>Contradiction explanation: Candesartan is evaluated with different co-treatment in two studies (anthracycline in the first study vs. trastuzumab in the second study). The combined drugs have differ-ent mechanisms of cardiotoxicity that may affect the Candesartan’s treatment outcome in different ways. |
| Study design | Contradictions may arise from differences in trial method-ology, such as parallel vs. crossover design, superiority vs. non-inferiority frameworks, blinding sta-tus, comparator type (placebo vs. active control), or randomization procedures. | Study 1 (PMID 28077507)<br>Context: *... patients were categorized as having paroxysmal, persis-tent, or permanent AF (atrial fibrillation) patterns at randomiza-tion...*<br>Claim: *The efficacy and safety profile of* ***edoxaban*** *as compared with* ***warfarin*** *was consistent across the 3 patterns of* ***AF.***<br>Study 2 (PMID 34962979)<br>Context: *... Of 21105 patients with AF randomized to warfarin, edoxaban 60/30mg, or edoxaban 30/15mg...*<br>Claim: ***edoxaban*** *30/15mg is inferior (vs.* ***warfarin****) for stroke prevention in* ***AF*** *patients with PAD.*<br>Contradiction explanation: Study 1 evaluates edoxaban across all AF subtypes broadly, while Study 2 focuses specifically on AF patients with a co-morbidity, peripheral artery disease. The different patient stratification strategies may result in different study outcomes. |
| Outcome measures | Contradictions may arise when studies assess dif-ferent outcomes or evaluate the same outcome at different time points. | Study 1 (PMID 26852138)<br>Context: *... assessed emotion processing during the presentation of pleasant, neutral and unpleasant pictures...*<br>Claim: ***ADHD*** *patients show a reduced reactivity towards pleasant stimuli.* ***Methylphenidate (MPH)*** *did not normalize this dysfunc-tion.*<br>Study 2 (PMID 27734305)<br>Context: *... Resting state functional connectivity (RSFC) analysis revealed significantly reduced connectivity under MPH compared to placebo...*<br>Claim: ***MPH*** *is ‘normalising’ a higher RSFC in young people with* ***ADHD.***<br>Contradiction explanation: The first study assesses behavioral emo-tion processing during stimulus presentation, while the second study evaluates resting-state functional connectivity. These outcomes cap-ture different aspects of ADHD, one reflecting behavioral responses and the other reflecting brain-level connectivity. The differences in outcome definitions may lead to contradictory evidence. |
| Ambiguous referent | One claim lacks a clear specifica-tion of species, population, dosage and expo-sure duration, or route of adminis-tration, resulting in uncertainty about the basis of comparison. | Study 1 (PMID 26822749)<br>Context: *... maintenance dose 4 mg/kg per day ...*<br>Claim: *support the concept of neuroprotection with* ***phenytoin*** *in patients with acute* ***optic neuritis*** *at concentrations at which ...*<br>Study 2 (PMID 32212009)<br>Context: (no dosage information mentioned)<br>Claim: ***Phenytoin*** *is not probably safe and effective as neuroprotec-tion after acute* ***optic neuritis.***<br>Contradiction explanation: The lack of dosage information in the second study abstract leading to ambiguity in interpretation of the second claim, prevents direct comparison between the two claims. |

Building on this taxonomy, we leverage LLMs to identify the study-level factor differences that explain the contradictory findings of the claims. Because a contradiction may come from several study differences at once, we treat explanation as a multi-label task. Specifically, we design the prompt to instruct LLMs to: (1) analyze the full abstract context of each claim; and (2) assign one or more study-level factors that explain the discrepancy between the claims. The output is structured as a set of study-level factor labels. The detailed prompt is presented in Appendix B.3.

### 2.5 Ground Truth Annotation

To establish ground truth for evaluating **DrugCon**’s contradiction detection and explanation outputs, we apply **DrugCon** to a large RCT collection, annotate its predictions, and then exhaustively review the remaining claim pairs. The resulting annotated dataset is DrugCon-Bench.

#### Data collection

We collect all RCT abstracts published from 2016 to 2025 in the PubMed Central Open Access subset, identified using Cochrane’s sensitivity- and precision-maximizing RCT query (Glanville et al. 2020). We apply **DrugCon** with **GPT-4.1-nano** as the LLM backbone. We choose **GPT-4.1-nano** for its low inference cost at the scale of this study and its high recall with the counterfactual prompt (Section 3.4). High recall is preferred, since false positives can be removed by annotators. Over the full corpus, claim pairing yields 609,877 candidate claim pairs, of which **DrugCon** predicts 615 as contradictory. These predicted pairs involve 819 unique abstracts, which form the annotation scope of DrugCon-Bench. To capture contradictions missed by **GPT-4.1-nano**, we exhaustively review all remaining claim pairs within these abstracts.

#### Annotation of DrugCon predictions

Four annotators participated: a biomedical informatics expert (HK) and three PhD students trained in biomedical NLP (ML, JM, and SL). For each **DrugCon**-predicted pair, annotators were shown the two claims side by side with their full abstracts, the predicted contradiction label, and the predicted study-level factors (Figure 3). They made two decisions: (a) whether the pair is an apparent contradiction, and (b) which study-level factors from the taxonomy (Section 2.4) explain it. Detailed guidelines are provided in Appendix A. To calibrate annotators and measure inter-annotator agreement, all four annotators labeled a shared subset of 50 pairs over three stages, meeting after each stage to reconcile disagreements and refine the guidelines. We stopped after the third stage because agreement had reached a satisfactory level (Section 3.1). Agreement was measured with Gwet’s AC1 (Gwet 2008), which is robust to the skewed label distributions. The remaining pairs were divided equally among annotators.

**Fig. 3.**
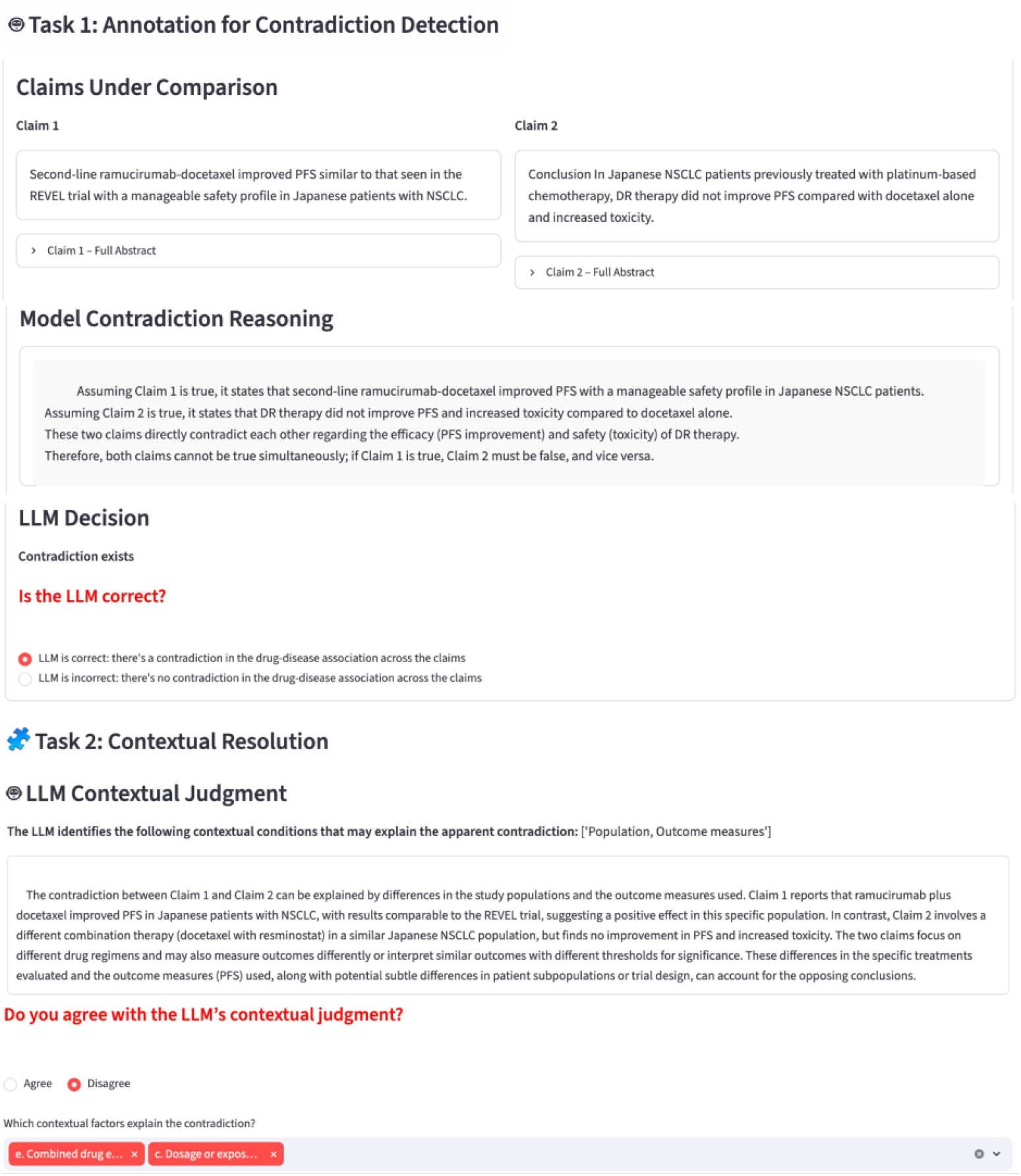
Screenshot of the annotation interface used for ground truth labeling of contradictory claim pairs by human annotators. Each screen displays the two claims, the full abstract contexts, and controls for labeling the contradiction identification and taxonomy-based explanation factors.

#### Exhaustive review

To capture contradictions that **DrugCon** missed, we review every remaining claim pair about the same drug–disease association within the 819 abstracts. One annotator (ML) identified 53 additional contradictory pairs, 48 of which were confirmed by a second annotator (HK). We do not attempt exhaustive annotation over all 224,756 abstracts, since the number of claim pairs grows quadratically with the number of abstracts; even after claim pairing, the full corpus yields 609,877 candidate pairs.

Because all contradictions within the 819 abstracts are annotated, DrugCon-Bench supports closed-world evaluation, unlike prior datasets that annotate only a subset of contradictions (Alamri and Stevenson 2016). Corpus statistics are reported in Section 3.1.

### 2.6 Evaluation

#### 2.6.1 Datasets

We evaluate on two datasets. DrugCon-Bench (Section 2.5) is our primary benchmark. To assess generalization, we also use ManConCorpus (Alamri and Stevenson 2016), a manually curated benchmark of contradictory claims from cardiovascular RCT abstracts. The two datasets share no abstracts, so ManConCorpus serves as an independent test of **DrugCon** method generalization.

ManConCorpus is originally structured as question–claim pairs, where each claim, extracted from RCT abstracts, either supports (“yes”) or refutes (“no”) a clinical question. To align this setting with our abstract-level contradiction detection task, and to better reflect real-world scenarios where claim annotations are not available for scientific abstracts, we convert the dataset into abstract-to-abstract pairs: two abstracts are labeled as non-contradictory if the pair of associated claims both support the same clinical question or both refute it, and contradictory otherwise.

We construct a question-based split, with 11 questions for training, 6 for development, and 8 for testing. This design ensures that all abstracts associated with a given clinical question are assigned to a single partition, preventing information leakage across splits and evaluating models’ generalization to unseen clinical topics at test time. All baseline models and the **DrugCon** framework are evaluated on the test set.

#### 2.6.2 Baselines

We compare **DrugCon**’s performance with the existing contradiction detection works: (1) a SemRep-based baseline, which we refer to as **SemRep-based method**, that detects contradictions by identifying pairs of subject–predicate–object triples sharing the same subject and object but exhibiting opposing predicates (e.g., *treats* vs. *causes*) (Rosemblat et al. 2019); (2) **DeBERTa-Base-Con**, a sentence-level classification model pretrained on the training part of ManConCorpus dataset (Alamri and Stevenson 2016) with DeBERTa-Base as the backbone model (Makhervaks et al. 2023) for sentence-level contradiction detection; (3) **MNLI-MedNLI**^1^, a natural language inference (NLI)–based model trained on MNLI (Williams et al. 2018) and MedNLI (Romanov and Shivade 2018), where contradiction detection is formulated as an entailment classification task (Deka et al. 2023).

The baseline models operate at the sentence-pair level, whereas our task is end-to-end contradiction detection from raw documents, including claim extraction, pairing, and contradiction classification. For a controlled comparison, we apply all baseline models to the candidate claim pairs generated in the second step of the **DrugCon** framework (Section 2.2), thereby holding the upstream steps fixed and isolating the contradiction classification component. Note that none of the baselines produces explanations. Contradiction explanation is therefore evaluated for **DrugCon** only.

#### 2.6.3 Metrics and Decoding Settings

For contradiction detection, we report precision, recall, and F_1_ for the contradictory class. For contradiction explanation, we report precision, recall, and F_1_ for each study-level factor, and micro-F_1_ across all factors. Explanation is evaluated on the 542 gold contradictory pairs in DrugCon-Bench.

Since the outputs of LLMs vary with the decoding temperature, we evaluate **DrugCon** under multiple decoding settings to ensure that its performance does not depend on a specific temperature. Specifically, we run **DrugCon** at temperatures 0, 0.5, and 1.0, and report the mean and standard deviation over the three runs. Similarly, we fine-tune **DeBERTa-Base-Con** with three random seeds and report the mean and standard deviation. In contrast, the **SemRep-based method** and **MNLI-MedNLI** are deterministic, as the **SemRep-based method** applies fixed rules over SemRep predications and **MNLI-MedNLI** performs inference with a fixed pretrained checkpoint, so neither involves any random component. We therefore run each of them once.

#### 2.6.4 Ablation Design

The **DrugCon** framework relies on two prompt-driven LLM components: contradiction *detection* (Section 2.3) and taxonomy-based contradiction *explanation* (Section 2.4). To understand the importance of backbone LLM selection and prompt design, we conduct an ablation study. We evaluate eleven backbone LLMs. Seven are open-weight models with known parameter counts, grouped into three size tiers: *≤*7B small models (**Ministral-3B**, **Gemma-3-4B**), 7– 20B medium models (**Llama-3.1-8B**, **Gemma-3-12B**), and *>*20B large models (**Gemma-3-27B**, **Qwen3-32B**, **Llama-3.3-70B**). The remaining four are proprietary models of undisclosed size: **Claude-Haiku-4.5**, **Claude-Sonnet-4.5**, **GPT-5.5**, and **GPT-4.1-nano** (the backbone used in our main experiments). For contradiction detection, to assess whether counterfactual reasoning helps uniformly regardless of the backbone model, we compare the *counterfactual* (CF) prompt, which asks whether one claim could still hold if the other is assumed true (Appendix B.1), against the *direct comparison* (DC) prompt, which skips the counterfactual reasoning and directly asks whether the two claims draw opposing conclusions about the drug’s effect on the disease (Appendix B.2). For explanation, we use the explanation prompt (Appendix B.3). All ablations use DrugCon-Bench at temperature 0, holding the upstream claim-extraction and claim-pairing stages fixed so that only the detection and explanation components vary.

## 3 Results

### 3.1 DrugCon-Bench Statistics

We collect 224,756 RCT abstracts published from 2016 to 2025; claim pairing yields 609,877 candidate claim pairs. As presented in Figure 4, **DrugCon** identified 615 candidate contradictory pairs from 819 abstracts, and annotators confirmed 494 of them. The exhaustive review found 48 more contradictory pairs. DrugCon-Bench therefore contains 542 (494+48) contradictory claim pairs from 819 abstracts. Inter-annotator agreement (Gwet’s AC1) in the final round was 0.812–0.839 (mean 0.821) for contradiction identification and 0.645–0.725 (mean 0.679) for factor labeling.

**Fig. 4.**
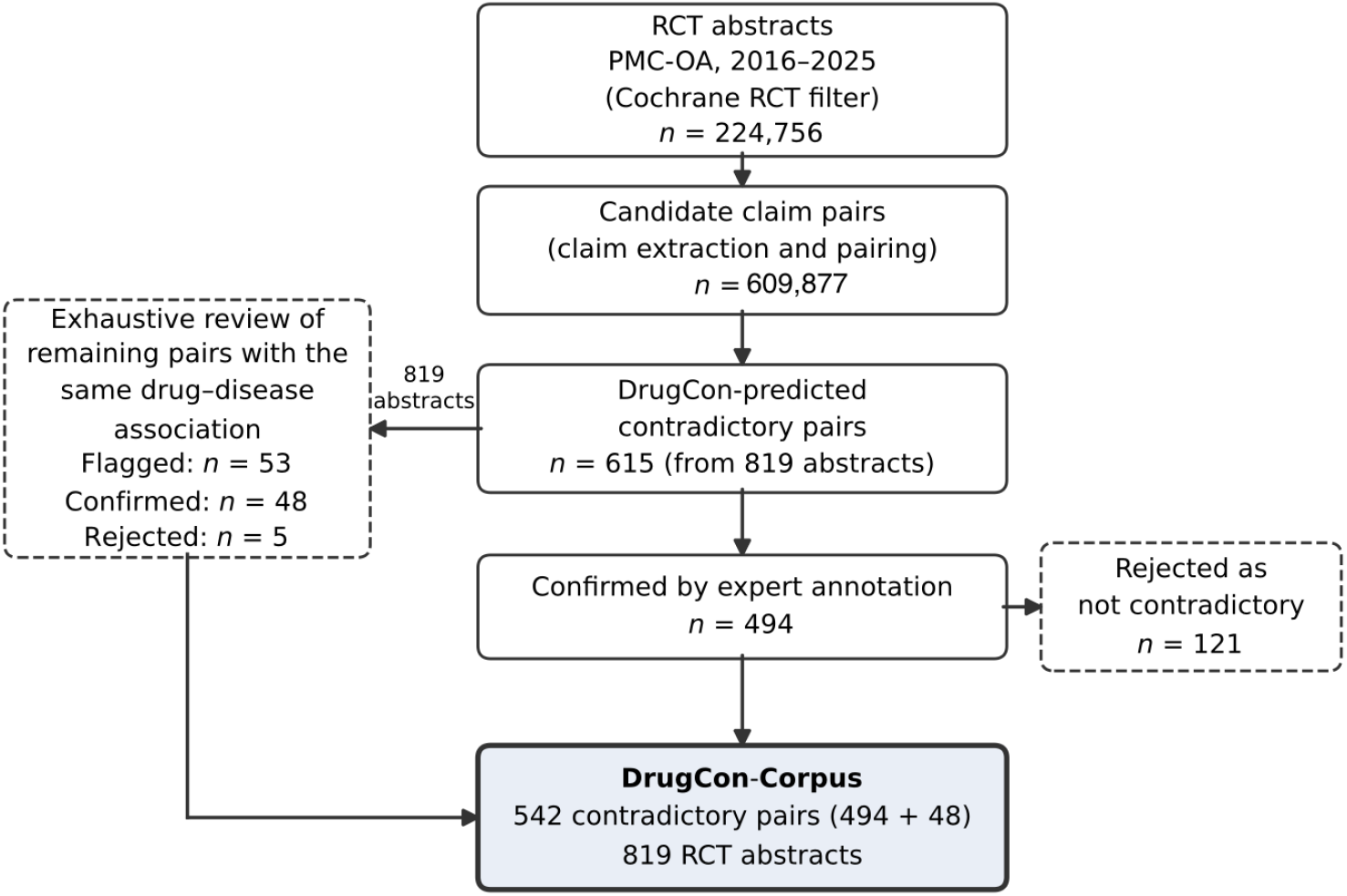
Construction of DrugCon-Bench. **GPT-4.1-nano** predicts 615 candidate contradictory pairs among 819 abstracts; annotators confirm 494, and an exhaustive review of the remaining claim pairs adds 48, yielding 542 contradictory pairs.

A contradiction can be explained by more than one contextual factor. In DrugCon-Bench, 75.5% of the contradictory pairs are annotated with two or more factors (2.18 factors per pair on average), so the percentages of factor distribution sum to more than 100%. The most common factors are Outcome measures (64.0%), Population (51.3%), and Dosage and exposure duration (35.6%), followed by Study design (20.3%), Combined drug effects (10.0%), and Route of administration (9.0%)(Figure 5).

**Fig. 5.**
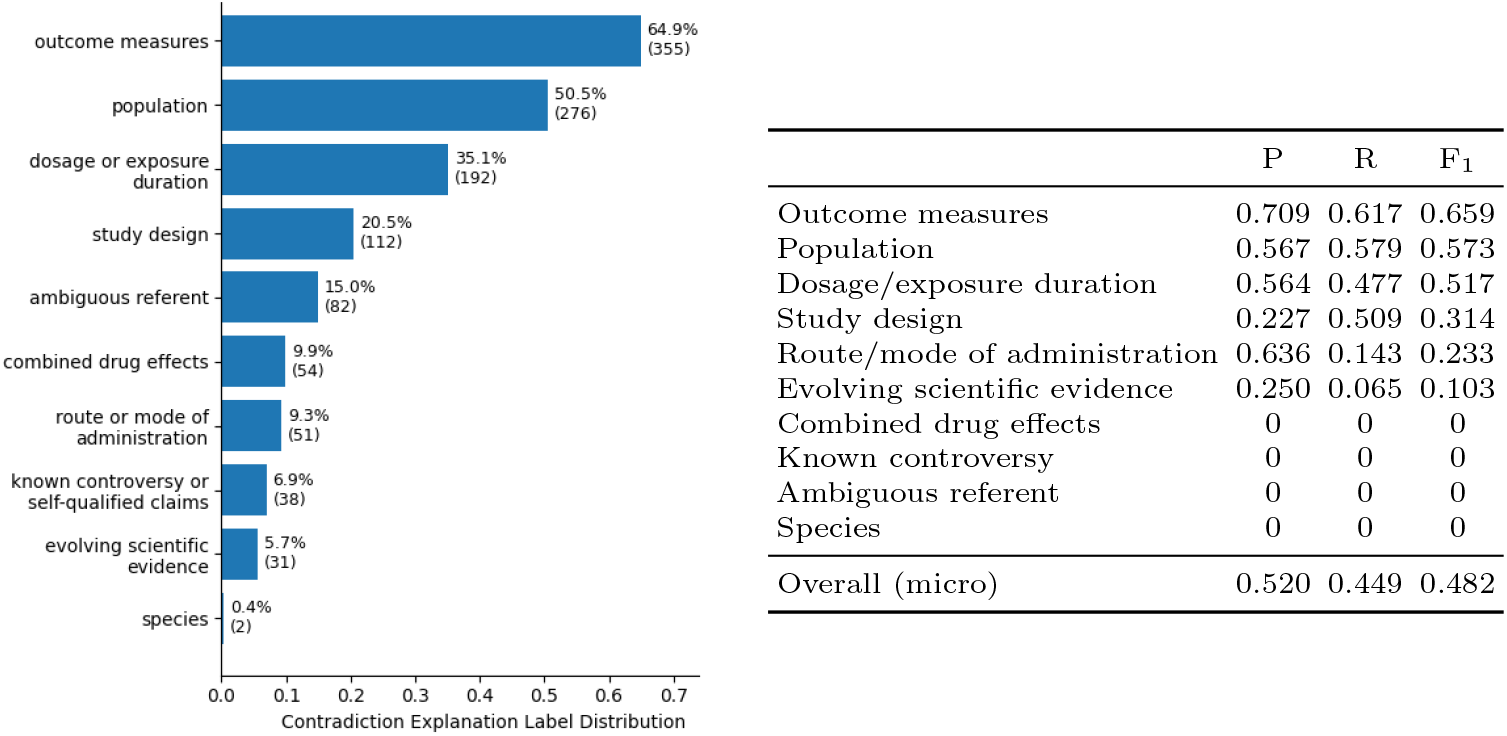
Left: distribution of study-level factors used to explain contradictions in DrugCon-Bench (human annotated). Note that percentages do not sum to 100% because a single contradiction sample may be associated with multiple factors. Right: performance of study-level contradiction explanation of **DrugCon** (backbone **GPT-4.1-nano**) from a single run at temperature 0. The performance on each study factor is presented.

### 3.2 Contradiction Detection Performance

#### Evaluation on DrugCon-Bench

Table 2 summarizes the performance of all methods on DrugCon-Bench. Note that the detection results in Table 2 come from separate runs at multiple decoding temperatures (Section 2.6.2), not from the run used to seed annotation. **DrugCon** significantly outperforms all baselines, with a precision of 0.636, recall of 0.896, and F_1_ score of 0.744. In contrast, all baseline methods exhibit lower performance:

- The rule-based **SemRep-based method** detects only 4 contradictions (2 correct), yielding a precision of 0.5 and extremely low recall (2/542). As discussed in its original work (Rosemblat et al. 2019), it captures apparent contradictions based on surface-level subject–predicate–object patterns, but fails to identify context-dependent or implicit contradictions, which are prevalent in DrugCon-Bench.
- The domain-specific model **DeBERTa-Base-Con**, trained on cardiology sentence pairs from ManConCorpus, does not generalize to DrugCon-Bench (F_1_=0.000), due to its cardiology focus.
- The NLI model, **MNLI-MedNLI**, identifies a small number of contradictions (recall: 0.106) but with very low precision (0.017). Despite being trained on the NLI corpus that covers general biomedical domain, it shows a tendency toward predicting “contradiction” for non-contradictory pairs in DrugCon-Bench.

**Table 2.** Contradiction detection performance across datasets. Standard deviations are reported for **DeBERTa-Base-Con** (three random seeds) and **DrugCon** (three decoding temperatures). **SemRep-based** and **MNLI-MedNLI** show no variation because they are deterministic. Best per dataset in **bold**. Asterisks on baseline F_1_ scores indicate that **DrugCon**’s F_1_ is statistically significantly higher than that of the baseline (Student’s *t*-test; *^∗^*: *p* < .05).

|  | DrugCon-Bench |  |  | ManConCorpus |  |  |
|---|---|---|---|---|---|---|
| Method | P | R | F<sub>1</sub> | P | R | F<sub>1</sub> |
| SemRep-based | 0.500 | 0.004 | 0.008* | 0.000 | 0.000 | 0.000* |
| DeBERTa-Base-Con | 0.000<sub>±0.0</sub> | 0.000<sub>±0.0</sub> | 0.000<sub>±0.0</sub>* | 0.436<sub>±0.034</sub> | 0.269<sub>±0.025</sub> | 0.333<sub>±0.029</sub>* |
| MNLI-MedNLI | 0.017 | 0.106 | 0.029* | 0.118 | 0.292 | 0.168* |
| **DrugCon** (ours) | **0.636**<sub>±0.035</sub> | **0.896**<sub>±0.026</sub> | **0.744**<sub>±0.028</sub> | **0.739**<sub>±0.148</sub> | **0.380**<sub>±0.112</sub> | **0.502**<sub>±0.129</sub> |

### Evaluation on ManConCorpus

Table 2 also reports contradiction detection performance on the ManConCorpus test set. The relative ordering of methods is consistent with our findings on DrugCon-Bench: **DrugCon** again achieves the best performance (F_1_ 0.502), significantly outperforming **DeBERTa-Base-Con** (0.333) and **MNLI-MedNLI** (0.168), with **SemRep-based method** again failing to detect any contradictions. The main difference is that **DrugCon** scores lower on ManConCorpus than on DrugCon-Bench (F_1_ 0.502 vs. 0.744), driven by lower recall (0.380 vs. 0.896), whereas **DeBERTa-Base-Con** and **MNLI-MedNLI** achieve higher recall on ManConCorpus (0.269 and 0.292) than on DrugCon-Bench (0.000 and 0.106).

### 3.3 Taxonomy-Based Explanation of Contradictions

We further evaluate how well **DrugCon** identifies the study-level factors that explain contradictions using our proposed taxonomy. Results are shown in Figure 5 (right). **DrugCon** attains an overall micro F_1_ of 0.482, where performance varies across factors. It performs best on the frequently occurring factors that are typically stated explicitly in RCT abstracts, including *Outcome measures* (F_1_=0.659), *Population* (F_1_=0.573), and *Dosage/exposure duration* (F_1_=0.517), indicating that the framework performs best on the dominant sources of disagreement. The moderate overall score reflects lower performance on rarer and more implicit factors, which we analyze below.

### 3.4 Ablation Study: Backbone LLM and Prompt Design

#### Contradiction detection

Table 3 reports detection performance for all eleven backbones under both prompts. Larger models perform better with the direct comparison prompt, which gives the best F_1_ for **Qwen3-32B** (0.737), **Llama-3.3-70B** (0.723), and three proprietary models (up to 0.793 for **Claude-Sonnet-4.5**). Smaller models instead perform better with the counterfactual prompt, including **Gemma-3-12B** (0.705 vs. 0.548), **Ministral-3B** (0.650 vs. 0.616), **Gemma-3-4B** (0.593 vs. 0.523), **Llama-3.1-8B** (0.583 vs. 0.554), and **GPT-4.1-nano** (0.744 vs. 0.649; OpenAI’s smallest and lowest-cost GPT-4.1 variant), as does **Gemma-3-27B** (0.678 vs. 0.635). Under the direct comparison prompt, these smaller models over-predict contradictions, achieving very high recall but low precision (e.g., **Gemma-3-12B** reaches 0.996 recall at 0.378 precision). Larger models are substantially more expensive: at the scale of 224,756 abstracts and 609,877 candidate pairs, running detection with **GPT-5.5** or **Claude-Sonnet-4.5** would cost approximately $5,500 and $2,800, respectively, compared with approximately $80 for **GPT-4.1-nano.**

**Table 3.** Ablation of backbone LLM and prompt design on DrugCon-Bench. For *detection*, we report precision (P), recall (R), and F_1_ under the counterfactual (CF, Appendix B.1) and direct comparison (DC, Appendix B.2) prompts. For *explanation*, we report multi-label micro-F_1_ under the explanation prompt (Appendix B.3). Models are grouped into open-weight size tiers and a proprietary group of undisclosed size. **GPT-4.1-nano** is the backbone used to construct DrugCon-Bench. The higher detection F_1_ between the two prompts is in **bold**. For **GPT-4.1-nano** detection, we report the mean over decoding temperatures from Table 2.

|  | Detection – CF |  |  | Detection – DC |  |  | Explanation |
|---|---|---|---|---|---|---|---|
| Model | P | R | F<sub>1</sub> | P | R | F<sub>1</sub> | Micro-F<sub>1</sub> |
| *≤7B* |  |  |  |  |  |  |  |
| Ministral-3B | 0.546 | 0.804 | **0.650** | 0.452 | 0.967 | 0.616 | 0.189 |
| Gemma-3-4B | 0.443 | 0.896 | **0.593** | 0.356 | 0.982 | 0.523 | 0.348 |
| *7–20B* |  |  |  |  |  |  |  |
| Llama-3.1-8B | 0.435 | 0.887 | **0.583** | 0.393 | 0.940 | 0.554 | 0.299 |
| Gemma-3-12B | 0.610 | 0.835 | **0.705** | 0.378 | 0.996 | 0.548 | 0.524 |
| *>20B* |  |  |  |  |  |  |  |
| Gemma-3-27B | 0.718 | 0.642 | **0.678** | 0.470 | 0.980 | 0.635 | 0.520 |
| Qwen3-32B | 0.856 | 0.501 | 0.632 | 0.633 | 0.883 | **0.737** | 0.567 |
| Llama-3.3-70B | 0.870 | 0.464 | 0.605 | 0.595 | 0.921 | **0.723** | 0.533 |
| *Proprietary (undisclosed size)* |  |  |  |  |  |  |  |
| Claude-Haiku-4.5 | 0.774 | 0.665 | 0.716 | 0.664 | 0.916 | **0.770** | 0.562 |
| Claude-Sonnet-4.5 | 0.800 | 0.667 | 0.728 | 0.717 | 0.887 | **0.793** | 0.590 |
| GPT-5.5 | 0.872 | 0.261 | 0.402 | 0.701 | 0.868 | **0.776** | **0.603** |
| GPT-4.1-nano | 0.636 | 0.896 | **0.744** | 0.494 | 0.945 | 0.649 | 0.482 |

#### Contradiction explanation

We further analyze taxonomy-based explanation performance across LLM backbones using the explanation prompt (Appendix B.3), reported in Table 3. The best micro-F_1_ increases from the *≤*7B tier (F_1_=0.348) to the 7–20B tier (F_1_=0.524), the *>*20B tier (F_1_=0.567), and the proprietary models (F_1_=0.603 for **GPT-5.5**). The smallest models perform worst, as **Ministral-3B** and **Llama-3.1-8B** both fall below 0.30 micro-F_1_.

## 4 Discussion

We presented **DrugCon**, an LLM-based framework for large-scale detection and structured explanation of contradictory drug–disease associations in RCT literature. Across both DrugCon-Bench and the external ManConCorpus benchmark, **DrugCon** with the **GPT-4.1-nano** backbone outperformed rule-based, supervised, and NLI-based baselines for contradiction detection. Beyond detection, **DrugCon** provides structured explanations based on study-level factors: it is effective at identifying differences in frequently occurring and explicit study factors, such as outcome measures, population, and dosage, but struggles with more implicit factors, such as evolving scientific evidence and ambiguous referents. These findings suggest that LLM-based reasoning is effective for large-scale contradiction analysis, while challenges remain in reasoning over scientific context and resolving implicit study factors.

### 4.1 Contradiction Detection Requires Implicit Reasoning

Existing approaches rely on lexical opposition and sentence-level information (Kim et al. 2006; Alamri 2016; Tawfik and Spruit 2018; Yazi et al. 2021; Makhervaks et al. 2023), which fail to capture implicit or cross-sentence knowledge. Our results suggest that contradiction detection in DrugCon-Bench instead requires implicit reasoning over the scientific claims. Many contradictory pairs do not contain explicit opposing statements (e.g., “effective” versus “ineffective”), but instead express disagreement through conditional statements or indirect assertions. Determining whether two claims are contradictory therefore requires interpreting the meaning and scope of the claims rather than relying on surface lexical cues or predicate oppositions. **DrugCon** addresses this by using an LLM to reason over the semantic implications of scientific claims and to integrate contextual information across multiple sentences. The following contradictory pair, which **DrugCon** identified but the baseline models missed, illustrates a contradiction that requires reasoning beyond explicit opposing statements:

Claim 1: *“Exercise was effective in reducing* ***fall-related injuries*** *among community-dwelling older women at a moderate cost.* ***Vitamin D supplementation had marginal additional benefit****.”* (PMID 26205890)

Claim 2: *“Our data suggest that* ***vitamin D supplementation*** *positively affected the rehabilitation program in patients [… ] by* ***reducing the risk of falls*** *and improving postural stability.”* (PMID 40220053)

The contradiction appears only when the two sentences of Claim 1 are read together: the “additional benefit” of vitamin D refers to the fall-related injuries mentioned in the previous sentence. Claim 1 thus indicates that vitamin D adds little to fall prevention, whereas Claim 2 reports that it reduces the risk of falls. Detecting this conflict requires cross-sentence reasoning and interpreting “marginal additional benefit” as a negative finding, rather than matching opposing words.

The lower recall of **DrugCon** on ManConCorpus can be mainly attributed to the definition of contradiction in ManConCorpus: each clinical question is paired with several claims that answer “Yes” or “No” to it, rather than with claims aligned on a specific relation. The following sample illustrates this:

Question: *“In patients with **coronary artery disease**, is mutation or polymorphisms in endothelial nitric oxide synthase gene associated with CAD or MI or ACS development?”*

Claim 1: *“We conclude that the E/D298 **polymorphism** is most consistently associated with **CAD**, but not with progression of atherosclerosis”* (PMID 15068395, labeled “yes”)

Claim 2: *“These findings suggest that the 894G>T, -786T>C and 4a/4b* ***polymorphisms*** *of the NOS3 were not associated with* ***CAD*** *in the studied subjects.”* (PMID 19931521, labeled “no”)

Because the two claims answer the question differently (“yes” vs. “no”), this pair is labeled as contradictory under our conversion of ManConCorpus (Section 2.6.1), while **DrugCon** labels it as non-contradictory. The two claims appear to examine different polymorphisms of the same gene (NOS3). Therefore, whether they are contradictory depends on the level of comparison. The claims may be contradictory at the gene level but compatible at the level of polymorphism each study examines. This ambiguity arises because ManConCorpus links claims through a shared high-level question rather than an explicitly aligned relation.

This comparison highlights another important contribution of this work: the construction of DrugCon-Bench. DrugCon-Bench reduces the ambiguity above by grounding each pair in an aligned drug–disease association and annotating contradictions directly at the claim level. It also provides a closed evaluation environment in which all pairwise contradictions within a subset of abstracts are exhaustively annotated, addressing a key limitation of the prior dataset (Alamri and Stevenson 2016), which provides only partial annotations of contradictory evidence. These properties support more precise and rigorous evaluation of contradiction detection systems in biomedical text.

### 4.2 From Detection to Explanation of Contradictions

A key contribution of this work is the extension from contradiction detection to contradiction explanation by study-level differences. Existing studies operate at the sentence level and treat contradiction detection as a binary task (Kim et al. 2006; Alamri 2016; Tawfik and Spruit 2018; Yazi et al. 2021; Makhervaks et al. 2023). **DrugCon** not only enables multi-sentence reasoning using LLMs, but also extends contradiction detection to structured explanation: it leverages the full abstract context to identify differences in study-level factors between two contradictory claims, such as population, dosage, and outcome measures, which may explain their different findings. **DrugCon** performs best on the factors that are stated explicitly in RCT abstracts, including Outcome measures (*F*_1_=0.659), Population (*F*_1_=0.573), and Dosage/exposure duration (*F*_1_=0.517).

However, **DrugCon** shows lower performance on the following relatively implicit factors:

- *Evolving scientific evidence* (F1=0.103) is annotated when conflicting claims reflect different stages of scientific understanding. As illustrated by the *eplerenone*–*heart failure* pair in Table 1, the earlier study (PMID 33175088) reports an inconclusive finding due to inadequate statistical power, while the later study (PMID 37370197) reports significant clinical benefits. Identifying this factor requires recognizing different time points of two studies. However, the current prompt design directs the model to focus solely on the abstract, which limits its ability to perform such temporal reasoning. Determining how two articles relate to one another, for example whether a later trial was designed to confirm, extend, or overturn an earlier one, requires information beyond the abstract, such as publication dates, citation links, shared trial registrations, and funding sources. Modeling such inter-article relationships needs further exploration in the future works.
- *Combined drug effects* (F1=0.000) is annotated for difference in co-administrations. As illustrated by the *candesartan*-*left ventricular function* pair in Table 1, candesartan is evaluated in combination with anthracycline in one study and trastuzumab in another. However, the LLM’s output for this pair labels the contradiction as *Study design*, explaining that the two studies differ in their trial designs. Although the LLM generated reasoning acknowledges the difference in co-administered drugs, the LLM attributes the difference to broader *study design* difference rather than to *Combined drug effects*. This suggests **DrugCon**’s low performance on *Combined drug effects* stems from the model attributing the co-administration situation to a more general category.
- For *Known controversy* (F1=0.000), **DrugCon** fails to identify any cases. This factor is annotated based on lexical cues, such as direct acknowledgment of further analysis requirement, uncertainty, or inconclusiveness within a claim, rather than differences between studies. As illustrated by the *melatonin*-*preoperative anxiety* pair in Table 1, Study 2 (PMID 37953202) explicitly acknowledges that early termination of the trial increases the likelihood of bias. However, the LLM’s generated reasoning over the contexts of this pair does not identify the self-acknowledged uncertainty of the claim 2. This suggests the prompt, currently designed to compare study factor differences between the study contexts described in the two abstracts, requires a further reasoning step for checking uncertainty markers within a single claim first.
- For *Ambiguous referent*, which is annotated when one claim lacks explicit contextual information such as dosage or population, **DrugCon** fails to identify any cases (F1=0.000). This failure stems from the prompt design rather than a fundamental limitation of LLMs. The current prompt instructs the model to assign the study-level factors that explain the contradiction based on differences between the two abstracts, so it is never asked to assess whether a factor is absent from an abstract. This suggests that explanation should be reframed as explicit extraction of each study-level factor, where “not reported” is a valid value, so that missing contextual information is identified directly rather than inferred. Such a structured extraction step could be further supported by checking the reporting completeness of each factor against established checklist items (Jiang et al. 2024, 2025).

Stronger backbones improve overall explanation performance (micro-*F*_1_ up to 0.603 for **GPT-5.5**; Table 3), but the implicit, low-support factors remain the hardest: even the best backbone attains only *F*_1_=0.436 on *evolving scientific evidence* and 0.022 on *ambiguous referents*. This is consistent with the error analysis above, as these failures arise from missing information and prompt design rather than from model capability alone.

### 4.3 Backbone and Prompt Selection

The ablation results (Section 3.4) show that no single prompt is best for every backbone, so prompt design and model choice should be considered together. Under the direct comparison prompt, smaller models over-predict contradictions, likely because judging whether two claims contradict in a single step provides little structure to guide a weaker model’s reasoning. The counterfactual prompt mitigates this problem by requiring an explicit, step-by-step check of whether the two claims can co-exist, which reduces over-prediction and improves precision (Wei et al. 2022). Larger models, in contrast, can more reliably judge whether two claims contradict in a single step, so the explicit reasoning steps of the counterfactual prompt provide little additional benefit. These stronger backbones, however, would cost roughly 35–70 times more than **GPT-4.1-nano** at the scale of this study. For explanation, performance increases with model capability, suggesting that identifying study-level factors, particularly the rarer and more implicit ones, requires deeper reasoning over the full abstract context.

### 4.4 Implications for Evidence-Based Medicine

Contradictory claims across RCTs present a challenge for evidence-based medicine (Prasad et al. 2013). Medical practitioners, clinicians, and clinical guideline developers should be careful when retrieving scientific findings to support their decisions. Ideally, all relevant findings, including contradictory results, should be considered to inform a more complete understanding of the evidence. **DrugCon** provides a scalable mechanism for identifying contradictory evidence and surfacing study-level factors that may explain these disagreements.

In clinical decision-making, by explicitly highlighting differences in populations, drug administration, or outcome definitions, **DrugCon** surfaces structured context that clinicians would otherwise need to identify manually across multiple studies. As discussed in the sample for *Population* in Table 1, when two studies report contradictory findings on effectiveness of indomethacin to treat post-ERCP pancreatitis, **DrugCon** highlights that the studies were conducted in different patient populations. This enables clinicians to assess which evidence is more applicable to their specific patient group. By making such study-level differences explicit, **DrugCon** could support more informed and nuanced interpretation of clinical findings.

**DrugCon** supports evidence synthesis by explicitly identifying the contradictory findings and the study-level differences underlying the contradictions. Firstly, contradictions across biomedical publications can hinder understanding of the current state of evidence, which challenges evidence review. The automated identification of contradictory claims therefore is important to support systematic reviewers and literature surveillance systems by highlighting findings that require closer examination (Ioannidis 2016; Kilicoglu 2018). Additionally, the identified study-level differences support the meta-analyses to capture the study-level variations when aggregating the results of multiple independent studies that address the same research question (Choi and Kang 2025).

For knowledge graph construction, **DrugCon** can support the development of more context-aware representations of biomedical knowledge. Existing biomedical knowledge graphs often aggregate findings from multiple publications into unified subject-predicate-object relations (Kilicoglu et al. 2012), where contradictory findings may coexist without explicit contextual information. When two contradictory claims coexist in the knowledge graph, it is not always clear which claim should be trusted (Sosa and Altman 2022). Current approaches to managing such contradictions rely on confidence scoring by source weighting (Wang et al. 2023), which may result in information loss by collapsing multiple context and metadata into a single score. The alternative approach could be representing claims together with their contextual characteristics, such as patient populations, outcome measures, and study design (Rosemblat et al. 2019). Rather than resolving contradictions by discarding or down-weighting claims, this preserves the full evidential context. By linking claims to their study-level factors, **DrugCon** preserves contextual information with the knowledge. This supports the construction of publication-specific or claim-centered knowledge graphs, enabling users to assess not only whether a relationship has been reported, but also under what conditions it was observed and whether contradictory evidence exists.

The large-scale contradiction analysis by **DrugCon** is also important for biomedical knowledge processing. Over 40 millions biomedical publications has been indexed in PubMed (Jin et al. 2024), which makes it practically impossible for clinicians and researchers to manually track the contradictory evidence across literature. **DrugCon** provides the scalable solution to processing this large-scale publications to support the downstream application of knowledge synthesis for clinicians, systematic reviewers, and knowledge graph developers.

### 4.5 Limitations and Future Work

Our study has several limitations. First, **DrugCon** is evaluated on RCT abstracts only. Although abstracts typically summarize the key findings and study characteristics (Schulz et al. 2010), they might omit details that might be relevant for explaining contradictory findings. Second, the **DrugCon** pipeline achieved an overall F1 score of 0.482 on the study-level factor difference identification. While the framework effectively identifies frequent factors such as outcome measures and population differences, it fails to identify combined drug effects, known controversy, and ambiguous referents, and rarely identifies evolving scientific evidence. Third, our contradiction taxonomy, adapted and extended from the previous work (Rosemblat et al. 2019), focuses on ten study-level factors. Although these factors capture many common sources of disagreement, they may not fully represent all reasons underlying contradictory findings.

## 5 Conclusion

Applying **DrugCon** to 224,756 RCT abstracts, we constructed DrugCon-Bench, a benchmark of exhaustively annotated contradictory claim pairs that supports closed-world evaluation of both contradiction detection and explanation. Across DrugCon-Bench and the external ManConCorpus benchmark, **DrugCon** outperforms rule-based, supervised, and NLI-based baselines, and moves beyond detection to identify the study-level factors that may explain why studies disagree. These results show that LLM-based reasoning is effective for large-scale contradiction analysis, while interpreting more implicit sources of disagreement remains challenging.

In future work, we will incorporate inter-article information (e.g., publication dates, citation links, and trial registrations) and reframe explanation as explicit extraction of each study-level factor (Section 4.2). We also plan to leverage the links between claims and their study-level factors to construct claim-centered knowledge graphs and evidence synthesis tools, enabling users to assess not only whether a drug-disease association has been reported, but also under what conditions it was observed and whether contradictory evidence exists.

## Declarations

## Acknowledgements

This work was partially supported by the National Library of Medicine of the National Institutes of Health, USA under the award number R01LM014079. The content is solely the responsibility of the authors and does not necessarily represent the official views of the National Institutes of Health. The funder had no role in considering the study design or in the collection, analysis, interpretation of data, writing of the report, or decision to submit the article for publication.

This research was partly supported by the Intramural Research Program of the National Institutes of Health (ZL). The contributions of the NIH author(s) are considered Works of the United States Government. The findings and conclusions presented in this paper are those of the author(s) and do not necessarily reflect the views of the NIH or the U.S. Department of Health and Human Services.

## Author Contributions

L.M.: Methodology, Data curation, Software, Validation, Formal analysis, Investigation, Writing – Original draft, Writing – Review & Editing. M.J.: Conceptualization, Data curation, Validation, Writing – Review & Editing. L.S.: Conceptualization, Data curation, Writing – Review & Editing. L.Z.: Conceptualization, Methodology, Data curation, Supervision, Project administration, Funding acquisition, Writing – Review & Editing. H.K.: Conceptualization, Methodology, Investigation, Supervision, Project administration, Funding acquisition, Writing – Original draft, Writing – Review & Editing. All authors have read and approved the manuscript.

## Data Availability

The RCT abstracts used in this study were obtained from the PubMed Central Open Access subset (https://pmc.ncbi.nlm.nih.gov/tools/openftlist/). DrugCon-Bench, including the 542 annotated contradictory claim pairs, the study-level factor labels, and the PMIDs of the 819 source abstracts, is publicly available at https://github.com/MengfeiLan/DrugCon_Drug_Contradiction/tree/main/DrugCon/data. ManConCorpus is available from its original publication (Alamri and Stevenson 2016).

## Code Availability

The source code of **DrugCon**, including claim extraction, claim pairing, contradiction detection and explanation prompts, and evaluation scripts, is publicly available at https://github.com/MengfeiLan/DrugCon_Drug_Contradiction/tree/main/DrugCon.

### Appendix A Annotation Guideline

We develop the online tool to support the annotation process. To complete the annotation process, please follow the instruction:

### 1. Introduction

Randomized controlled trials (RCTs) are the gold standard of biomedical evidence, while their findings can sometimes be contradictory. Drug–disease associations are especially important because they directly guide clinical decisions, inform drug repurposing efforts, and support health policy. Contradictory findings across studies can create confusion, hinder reliable evidence synthesis, and slow down the translation of research into practice. It is important to develop automated systems that can systematically detect and resolve contradictory drug–disease claims, providing clinicians and researchers with a clearer, more reliable evidence base.

To address this, we developed an LLM-based framework to identify potential contradictions across large-scale RCT articles. The framework consists of:

- Claim Extraction: extracting scientific claims from RCT abstracts.
- Related Claim Pairing: pairing semantically related claims using rule-based methods and dense embeddings.
- Contradiction Detection: using LLM prompting to determine whether claim pairs are contradictory.

We collected 224,756 RCT abstracts (2016-2025). The framework identified 615 potential drug–disease contradictions requiring human validation.

### 2. Annotation Tasks

**Task 1: Binary contradiction verification.** Determine whether you agree with the model’s judgment that two claims are contradictory.

*Key instructions:*

- Use only the two claim sentences.
- Focus on drug–disease relationship
- Resolve abbreviations using context.

**Task 2: Contextual resolution.** Determine whether contradictions can be explained by contextual factors. If so, select applicable taxonomy categories.

### 3. Contextual Factor Taxonomy

#### Species

Differences across biological species (e.g., animal vs human studies).

#### Population

Stable participant characteristics defining study groups. Decision rule: differences in who qualifies for inclusion.

Examples: age, sex, ethnicity, pregnancy, disease type/severity, comorbidities.

#### Dosage and exposure duration

Amount and duration of intervention (dose, frequency, length of treatment).

#### Route or mode of administration

How intervention is delivered (oral, IV, inhalation, timing differences).

#### Evolving scientific evidence

Contradictions due to updated scientific understanding or revised hypotheses.

#### Known controversy

Explicit uncertainty or acknowledged disagreement in claims.

#### Combined drug effect

Drug–drug interactions or co-administration effects.

#### Study design

Differences in trial structure (randomization, blinding, crossover, parallel, analysis type).

#### Outcome measures

Different endpoints or different measurement time points.

#### Ambiguous referent

One claim lacks explicit contextual specification (dose, population, route, species) while the other provides it.

#### Other

Unclassified factors.

### Appendix B Prompt Templates

#### B.1 Contradiction Detection Prompt - Counterfactual Reasoning

###### Task

Decide whether two biomedical claims about the same drug–disease association present a contradiction, using counterfactual reasoning.

(1) Assume Claim 1 is true. Under this assumption, would Claim 2’s overall assessment of the drug for the disease still hold?

(2) Assume Claim 2 is true. Under this assumption, would Claim 1’s overall assessment of the drug for the disease still hold?

###### Definition

A “contradiction” here is an apparent contradiction: the two claims give opposing overall assessments of the same drug for the same disease (e.g., effective vs. ineffective, beneficial vs. harmful, safe vs. unsafe). It need not be a strict logical impossibility.

###### Task Notes

(a) The two claims are contradictory if either (1) or (2) reveals that the overall assessments oppose each other.
(b) A contradiction may be partial: if one subclaim or subpoint in a claim opposes the other claim, treat the pair as contradictory, even if other parts agree.
(c) Do not dismiss a contradiction merely because the two studies differ in outcome measured, patient subgroup, dose, duration, timing, comparator, or study design. Opposing conclusions still count as a contradiction even if they could be reconciled by such differences.
(d) Classify as “Claims can coexist” only if the claims concern different drugs or diseases, express the same overall direction, or make no clear assessment of the drug–disease association.

**Claim 1:** [Claim 1 Content]

**Claim 2:** [Claim 2 Content]

**Output Format:** Return your answer in the exact structure below:

<result>

<reasoning> [Your reasoning steps here]

</reasoning>

<conclusion> [One of: "Contradiction exists", "Claims can coexist"]

</conclusion>

</result>

Using a simple rule-based parser, we extract the contradiction label for each claim pair directly from the <conclusion> block. Similarly, we capture the content within the <reasoning> block to analyze the model’s step-by-step justification, enabling subsequent analyses of its reasoning behavior.

#### B.2 Contradiction Detection Prompt - Direct Comparison

###### Task

Determine whether two biomedical claims contain a potential contradiction about the same drug–disease association. For this task, “contradiction” means a potential contradiction in the overall assessment of the drug for the disease, not a strict logical impossibility.

Focus only on:

- the drug, treatment, or intervention
- the disease, condition, or indication
- the overall direction/valence of the claim about that drug–disease association

Classify as “Contradiction exists” if the claims concern the same drug or drug class and the same disease/condition, and their overall assessments are discordant. Do not reject a contradiction merely because the claims differ in population, outcome, endpoint, biomarker, comparator, dose, duration, study design, or statistical method.

Classify as “Claims can coexist” only if the claims are about different drugs or diseases, share the same overall direction, or do not make a clear assessment of the drug–disease association.

###### Task Notes

Keep the reasoning short and direct. Identify the drug, the disease, and the overall direction of each claim.

**Claim 1:** [Claim 1 Content]

**Claim 2:** [Claim 2 Content]

**Output Format:** Return your answer in the exact structure below:

<result>

<reasoning> [Briefly identify the drug, disease, and overall

direction of each claim. State whether the directions are

discordant.]</reasoning>

<conclusion> [One of: "Contradiction exists", "Claims can coexist"]

</conclusion>

</result>

In contrast to the counterfactual prompt in Appendix B.1, which determines a contradiction through bidirectional “assume-true” reasoning, the direct comparison prompt asks the model to judge in a single step whether the two claims reach opposing conclusions about the same drug–disease association. We use a simple rule-based parser to extract the binary contradiction label from the <conclusion> block, as described in Appendix B.1.

#### B.3 Contradiction Explanation Prompt

###### Task

Given two biomedical claims that appear to report opposing outcomes for the same drug–disease association, analyze whether the apparent contradiction can be explained by differences in study-level contextual conditions.

Specifically: (1) Examine Claim 1 together with its source abstract. (2) Examine Claim 2 together with its source abstract. (3) Determine whether the opposing outcomes can be explained by one or more contextual factors from the taxonomy below, and provide an appropriate explanation of how each identified factor accounts for the contradiction.

###### Internal-to-patient factors

- **Species.** The claims are based on different species, where differences in disease manifestation, biological mechanisms, and pharmacokinetics across species limit the direct comparability of drug effects.
- **Population.** The claims target different human subpopulations (e.g., age, sex, genetics, comorbidities, ethnicity, or risk profiles).

###### External-to-patient factors

- **Dosage and exposure duration.** The same intervention is administered at different doses or exposure durations, which may produce different biological effects.
- **Route or mode of administration.** The intervention is delivered via different routes or timing, which may change tissue targeting or absorption.
- **Evolving scientific evidence.** The claims reflect different stages of scientific understanding, where early hypotheses are later revised by subsequent work.
- **Known controversy.** One or both claims explicitly acknowledge uncertainty, debate, or conditional validity, indicating an ongoing scientific controversy rather than a definitive conclusion.
- **Combined drug effect.** The reported effect of a drug depends on its combination with other medications, where drug–drug interactions or synergistic effects produce outcomes that differ across studies.
- **Study design.** The studies differ in trial methodology, such as parallel vs. crossover design, superiority vs. non-inferiority frameworks, blinding status, comparator type (placebo vs. active control), or randomization procedures.
- **Outcome measures.** The studies assess different outcomes or evaluate the same outcome at different time points.
- **Ambiguous referent.** One claim lacks a clear specification of species, population, dosage and exposure duration, or route of administration, resulting in uncertainty about the basis of comparison.

###### Task Notes

Base your analysis strictly on the provided claims and abstracts. Do not use external biomedical knowledge. Do not speculate beyond what is explicitly stated in the text.

**Claim 1:** [Claim 1 Content]

**Claim 1 context:** [Abstract 1]

**Claim 2:** [Claim 2 Content]

**Claim 2 context:** [Abstract 2]

**Output Format:** Return your answer in the exact structure below:

<result>

<conclusion>

contextual_factor: [comma-separated factor names | none]

</conclusion>

<explanation>

[Brief explanation describing how each identified contextual factor accounts for the apparent contradiction, with references to specific textual evidence.]

</explanation>

</RESULT>

### Appendix C Performance of Item-Level Contradiction Explanation

Table C1 presents the contradiction explanation breakdown by taxonomy items, one row per model and one column per contextual factor, ordered by gold support. First, the aggregate score is carried by the three high-support factors *outcome measures* (OM, *n*=347), *population* (POP, *n*=278), and *dosage/exposure duration* (DOS, *n*=193), on which the stronger backbones achieve F1 0.6–0.8; the remaining factors contribute little to the micro total. Secondly, the implicit, discourse-level factors stay hard: *ambiguous referent* (AMB, *n*=82) is zero for all models except **GPT-5.5** (0.022), and *known controversy* (KC, *n*=38) reaches 0.203 only for **GPT-5.5** and is near zero elsewhere, while *evolving scientific evidence* (ESE, *n*=31) is recovered mainly by the frontier models, including **GPT-5.5**, **Claude-Sonnet-4.5** and **Claude-Haiku-4.5**.

**Table C1.**
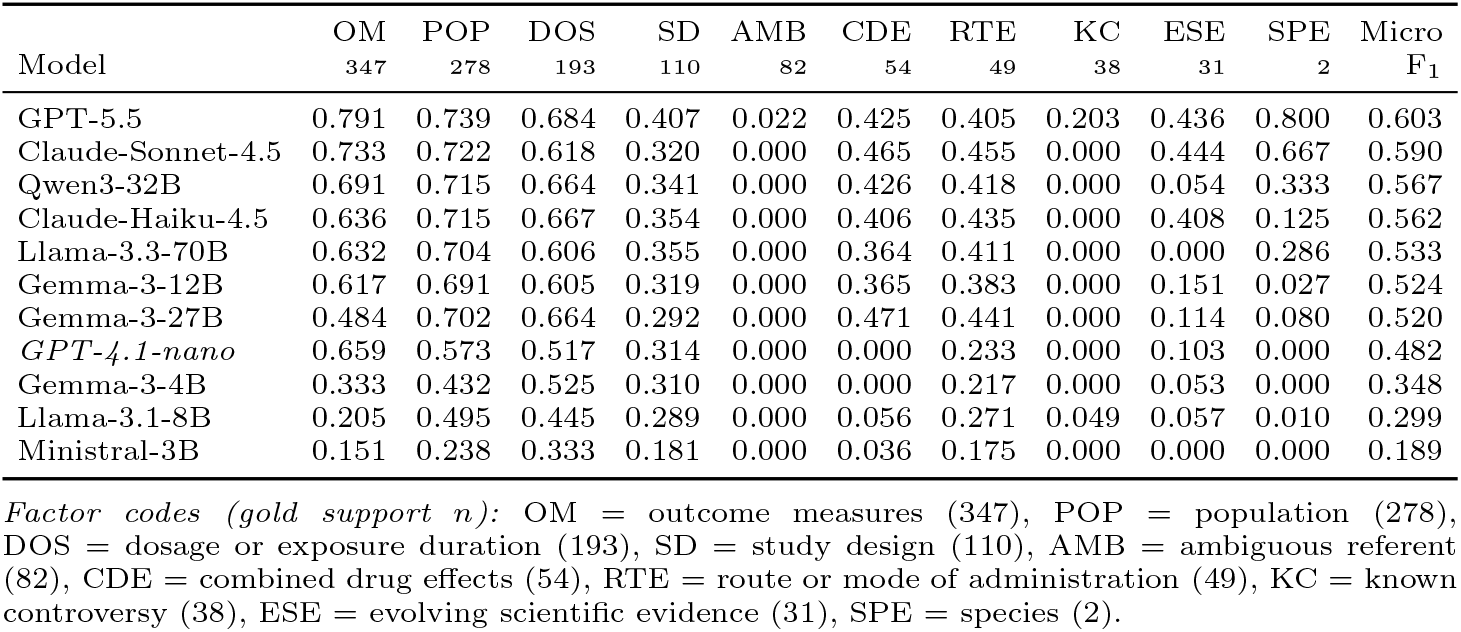
Per-factor contradiction explanation F_1_ of each backbone LLM on DrugCon-Bench, using the explanation prompt (Appendix B.3). Factor columns are ordered by gold support *n* (shown in the header). Rows are sorted by micro-F_1_ (descending). **GPT-4.1-nano** (*italic*) is the backbone used in the main experiments.

|  | OM | POP | DOS | SD | AMB | CDE | RTE | KC | ESE | SPE | Micro |
|---|---|---|---|---|---|---|---|---|---|---|---|
| Model | 347 | 278 | 193 | 110 | 82 | 54 | 49 | 38 | 31 | 2 | F<sub>1</sub> |
| GPT-5.5 | 0.791 | 0.739 | 0.684 | 0.407 | 0.022 | 0.425 | 0.405 | 0.203 | 0.436 | 0.800 | 0.603 |
| Claude-Sonnet-4.5 | 0.733 | 0.722 | 0.618 | 0.320 | 0.000 | 0.465 | 0.455 | 0.000 | 0.444 | 0.667 | 0.590 |
| Qwen3-32B | 0.691 | 0.715 | 0.664 | 0.341 | 0.000 | 0.426 | 0.418 | 0.000 | 0.054 | 0.333 | 0.567 |
| Claude-Haiku-4.5 | 0.636 | 0.715 | 0.667 | 0.354 | 0.000 | 0.406 | 0.435 | 0.000 | 0.408 | 0.125 | 0.562 |
| Llama-3.3-70B | 0.632 | 0.704 | 0.606 | 0.355 | 0.000 | 0.364 | 0.411 | 0.000 | 0.000 | 0.286 | 0.533 |
| Gemma-3-12B | 0.617 | 0.691 | 0.605 | 0.319 | 0.000 | 0.365 | 0.383 | 0.000 | 0.151 | 0.027 | 0.524 |
| Gemma-3-27B | 0.484 | 0.702 | 0.664 | 0.292 | 0.000 | 0.471 | 0.441 | 0.000 | 0.114 | 0.080 | 0.520 |
| *GPT-4.1-nano* | 0.659 | 0.573 | 0.517 | 0.314 | 0.000 | 0.000 | 0.233 | 0.000 | 0.103 | 0.000 | 0.482 |
| Gemma-3-4B | 0.333 | 0.432 | 0.525 | 0.310 | 0.000 | 0.000 | 0.217 | 0.000 | 0.053 | 0.000 | 0.348 |
| Llama-3.1-8B | 0.205 | 0.495 | 0.445 | 0.289 | 0.000 | 0.056 | 0.271 | 0.049 | 0.057 | 0.010 | 0.299 |
| Ministral-3B | 0.151 | 0.238 | 0.333 | 0.181 | 0.000 | 0.036 | 0.175 | 0.000 | 0.000 | 0.000 | 0.189 |

*Factor codes (gold support n):* OM = outcome measures (347), POP = population (278), DOS = dosage or exposure duration (193), SD = study design (110), AMB = ambiguous referent (82), CDE = combined drug effects (54), RTE = route or mode of administration (49), KC = known controversy (38), ESE = evolving scientific evidence (31), SPE = species (2).

## Footnotes

1 https://huggingface.co/pritamdeka/PubMedBERT-MNLI-MedNLI

